# Does malignancy status effect outcomes in patients with large vessel occlusion stroke and cancer that underwent endovascular thrombectomy?

**DOI:** 10.1101/2023.03.23.23287554

**Authors:** Fatma Shalabi, Tzvika Sacagiu, Asaf Honig, Jeremy Molad, Zeev Itsekson-Hayosh, Hen Hallevi, David Orion, Shorooq Aladin, John M. Gomori, Jose E. Cohen, Ronen R. Leker

**Affiliations:** Department of Neurology, Hadassah-Hebrew University Medical Center, Jerusalem, Israel; Department of Neurology, Tel Aviv Sourasaky Medical Center, Tel Aviv, Israel; Department of Neurology, Sheeba Medical Center Ramat Gan Israel; Department of Radiology, Hadassah-Hebrew University Medical Center, Jerusalem, Israel; Department of Neurosurgery, Hadassah-Hebrew University Medical Center, Jerusalem, Israel

**Keywords:** Cancer, Stroke, Thrombectomy

## Abstract

**Background:** Cancer is associated with an increased risk of acute ischemic stroke (AIS) including large vessel occlusions (LVO). Whether cancer status affects outcomes in patients with LVO that undergo endovascular thrombectomy (EVT) remains unknown.

**Methods and Results:** All consecutive patients undergoing EVT for LVO were recruited into a prospective ongoing multi-center database and the data was retrospectively analyzed. Patients with active cancer were compared to patients with cancer in remission. Association of cancer status with 90-day functional outcome and mortality were calculated in multivariable analyses. We identified 154 patients with cancer and LVO that underwent EVT (mean age 74±11, 43% men, median NIHSS 15). Of the included patients, 70 (46%) had a remote history of cancer or cancer in remission and 84 (54%) had active disease. Outcome data at 90 days post-stroke was available for 138 patients (90%) and was classified as favorable in 53 (38%). Patients with active cancer were younger and more often smoked but did not significantly differ from those without malignancy in other risk factors, stroke severity, stroke subtype or procedural variables. Favorable outcome rates among patients with active cancer did not significantly differ compared to those seen in patients without active cancer but mortality rates were significantly higher among patients with active cancer on univariate and multivariable analyses.

**Conclusions:** Our study suggests that EVT is safe and efficacious in patients with history of malignancy as well as in those with active cancer at the time of stroke onset although mortality rates are higher among patients with active cancer.

**Clinical Perspectives:** *What is new?:*

- The results of the current study suggest that EVT is safe and efficacious in patients with history of malignancy as well as in those with active cancer at the time of stroke onset although mortality rates are significantly higher and favorable outcome rates tend to be lower among patients with active cancer.

*What are the clinical implications?:*

- The results imply that all cancer patients with LVO stroke should be considered for EVT regardless of cancer status.

## Introduction

The prevalence of cerebrovascular disease in the setting of malignancy is approximately 15%.^1-3^ Cancer associated coagulopathy is the main pathological mechanism underlying cancer-related stroke (CRS).^1-3^ Other causes include radiation chemotherapy and biological therapy are all associated with potential vessel wall injury and coagulopathy.^4-9^ Additionally, many cancer patients are prone to have stroke secondary to concomitant atherosclerotic disease secondary to hypertension, diabetes mellitus, hyperlipidemia and smoking as well as to atrial fibrillation. Patients with stroke secondary to large vessel occlusion (LVO) have higher rates of recanalization and favorable outcomes when treated with endovascular thrombectomy (EVT).^10^ Several case series published lately suggest that EVT may be as effective in achieving target vessel recanalization in cancer patients with stroke and LVO although the chances for favorable outcome are somewhat reduced in cancer patients.^11-24^ However, only one of these studies differentiated between patients with active cancer to those with cancer in remission.^24^ Therefore, our goal was to determine whether cancer state at the time of stroke affects outcomes in cancer patients with LVO that underwent EVT.

## Methods

The data that support the findings of this study are available from the corresponding author upon reasonable request in compliance with governmental rules.

We conducted a retrospective analysis of our prospective multicenter database for all patients arriving with LVO that underwent EVT at three tertiary academic centers. Data from all patients undergoing EVT were accrued continuously without specific exclusion criteria. The study was approved at each individual center by the local institutional review boards at Hadassah Medical Organization, Tel Aviv Medical Center and Chaim Sheeba Medical Center and permissions to combine anonymized individual patient data into a unified dataset was granted with a waiver to obtain informed consent, given the retrospective nature of the data analysis.

We identified patients with a history of remote cancer in remission or suffering from an active malignancy at the time of presentation. These patients were identified based on personal history obtained using a standardized questionnaire administered to all patients and family members or from medical records when the questionnaire could not be filled (e.g. in aphasic patients with no family member present). We compared between patients with active cancer to those with history of cancer but no evidence of active disease. For the purpose of the current study, active cancer was defined as a diagnosis of cancer, other than local basal-cell carcinoma of the skin within 180 days prior to LVO, any treatment for cancer within 180 days prior to LVO, new diagnosis of cancer during admission for LVO or recurrent or metastatic cancer at the time of LVO.^25^

Clinically relevant accrued data included demographics, time metrics, baseline stroke severity determined with the National Institutes of Health Stroke Scale (NIHSS) and outcome status determined with the modified Rankin Scale (mRS) scores at discharge and 3 months after discharge. Radiological data included NCCT and/or magnetic resonance imaging (MRI) and CTA data acquired before the endovascular treatment. Treatment with intravenous tPA was given according to recommendations for thrombolytic treatment in the institutional guidelines. A history of cancer was not considered a contraindication for tPA unless the presence of brain metastasis was known or suspected or the patient was considered to present high risk of systemic bleeding related to the cancer or systemic metastasis.

Experienced interventional specialists treated all patients undergoing EVT. All procedures were performed using a femoral artery approach under general anesthesia. A balloon guide catheter was routinely used unless technically unfeasible, and a heparinized saline solution was continuously perfused through the catheter during the procedure. Mechanical thrombectomy using stent retrievers or aspiration techniques were attempted at the discretion of the endovascular specialists.

Data on procedural variables including the thrombolysis in cerebral infarction (TICI) score^26^ at the end of the procedure and the number of passes needed to achieve the best possible recanalization status were also studied. TICI2b-3 was considered as successful target vessel recanalization.

Patients were seen after discharge at the outpatient stroke clinics. We also obtained the information on mortality by review of the medical records.

Primary outcomes included functional outcome determined with the modified Rankin Scale (mRS) at 3-month post EVT. Favorable outcome was defined as either an mRS ≤2 at 90 days for patients that were independent prior to stroke and no change in mRS for those with pre-existing disability (mRS≥3) prior to the stroke.

Secondary safety outcomes were symptomatic hemorrhagic transformation (symptomatic intracranial hemorrhage, sICH) according to the ECASS criteria^27^ and death.

### Statistical analysis

Statistical analysis was performed using the SPSS 25 (IBM USA). p < 0.05 was considered significant. The □^2^ test was used to explore the link between qualitative variables. The student’s t-test was used to compare quantitative variables. We next performed multivariable logistic regression modeling to test impact on favorable outcome and survival. Only variables with p < 0.1 were entered into our regression analysis.

## Results

Among 1263 patients with acute LVO that underwent EVT and were included in the registry, 154 patients (43% men, median age 74 [IQR 67-82]) had a concomitant history of cancer (Figure 1). The most common cancer types included breast (21%), lung (14%), colon (11%) and lymphoma (6%). The median NIHSS was 15 (IQR 15-20) on admission and 5 (IQR 5-12) at discharge. Favorable target vessel recanalization (TICI≥2b) was achieved in 132 (85%) patients and the median number of passes needed to recanalize the vessel was 2 (IQR 1-3). sICH was observed in 10 patients (6.5%). Outcome data at 90 days post stroke was available for 139 (90%) patients with 15 patients lost to follow-up. Among patients with available follow up data, 54 (39%) achieved a favorable functional outcome and 44 (34%) were dead before 90 days from onset.

**Figure 1.**
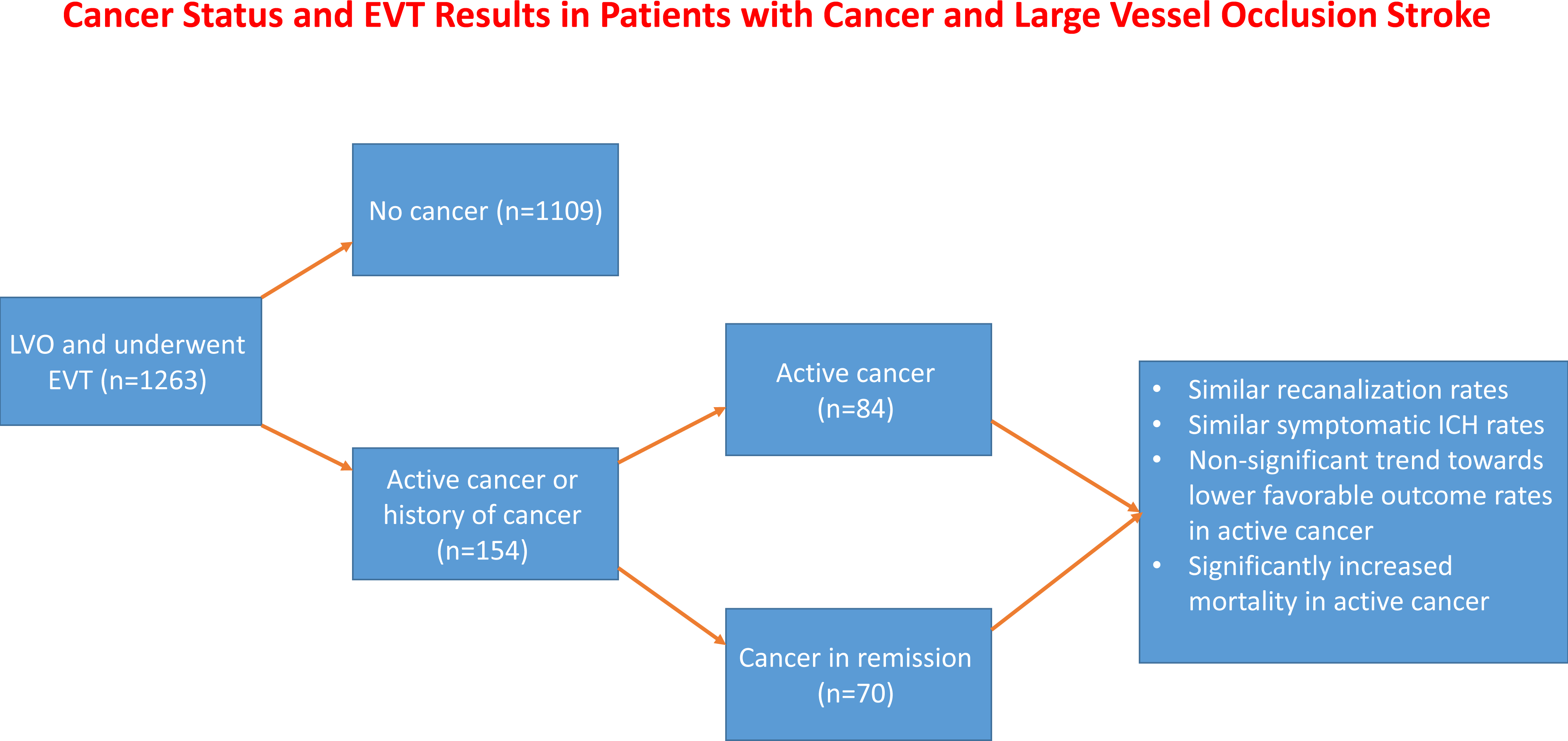
Cancer status and EVT results in patients with cancer and large vessel occlusion stroke. The Figure shows study flow chart and outcomes

Of the included cancer patients, 84 (54%) had an active malignancy at the time of presentation (mean age±SD 71.6±12.2, 51% men), while 70 (46%) had a history of cancer with no known activity at the time of EVT (mean age±SD, 75.5±11.1, 39% men). Comparison of patients with active cancer to those with a history of cancer (Table 1) showed that patients with active cancer were significantly younger and were more often smokers. However, other baseline risk factors as well as stroke severity, suspected stroke etiology and site of vessel occlusion did not differ between the groups. Similarly, the frequency of use of bridging with systemic thrombolysis did not differ between the groups. Procedural variables including time metrics and number of passes needed to achieve target vessel recanalization also did not significantly differ between the groups. Favourable target vessel recanalization defined as TICI2b-3 was achieved in 86% of patients in both groups. Only 10 patients were found to have symptomatic hemorrhagic transformations and there was no significant difference in the frequency of ICH between the groups.

**Table 1.** Baseline Characteristics of the Patients

|  | All (n=154) | Active cancer<br>N=84 | Cancer history<br>N=70 | p-value |
| --- | --- | --- | --- | --- |
| Age, (±SD) | 73.4±11.7 | 71.6±12.2 | 75.5±10.8 | 0.04 |
| Sex, male (%) | 71 (46%) | 43 (51%) | 28 (40%) | 0.16 |
| HTN (%) | 102 (66%) | 51 (61%) | 51 (73%) | 0.11 |
| DM (%) | 47 (31%) | 24 (29%) | 23 (33%) | 0.56 |
| Dyslipidemia (%) | 70 (46%) | 41 (49%) | 29 (41%) | 0.08 |
| AF (%) | 62 (40%) | 32 (38%) | 30 (43%) | 0.08 |
| Smoking (%) | 39 (25%) | 31 (37%) | 8 (11%) | 0.001 |
| IHD (%) | 62 (40%) | 32 (38%) | 30 (43%) | 0.08 |
| Prior stroke (%) | 27 (17.5%) | 13 (15.5%) | 14 (20%) | 0.46 |
| Antiplatelets | 31 (21%) | 19 (24%) | 12 (18%) | 0.38 |
| Anticoagulants | 31 (20%) | 18 (21%) | 13 (19%) | 0.66 |
| Vessel lesion (%) |  |  |  | 0.41 |
| ICA | 23 (15%) | 13 (15%) | 10 (14%) |  |
| MCA | 93 (60%) | 47 (56%) | 46 (66%) |  |
| Tandem | 8 (5%) | 5 (6%) | 3 (4%) |  |
| Other | 30 (20%) | 19 (23%) | 11 (16%) |  |
| TOAST (%) |  |  |  | 0.07 |
| Cardioembolic | 67 (44%) | 36 (43%) | 31 (44%) |  |
| Large vessel atherosclerosis | 57 (37%) | 26 (31%) | 31 (44%) |  |
| Other known | 6 (4%) | 5 (6%) | 1 (1%) |  |
| Unknown | 24 (16%) | 17 (20%) | 7 (10%) |  |
| Initial NIHSS score (±SD) | 15.1±6.0 | 14.6±6.3 | 15.6±5.5 | 0.30 |
| IV tPA (%) | 38 (25%) | 16 (19%) | 22 (31%) | 0.08 |
| Baseline mRS (%) |  |  |  | 0.32 |
| 0-2 | 125 (81%) | 66 (78%) | 59 (84%) |  |
| 3 | 23 (15%) | 14 (17%) | 9 (13%) |  |
| 4 | 6 (4%) | 4 (5%) | 2 (3%) |  |

mRS – modified Rankin Scale, NIHSS- National Institutes of Health Stroke Scale, TICI – thrombolysis in cerebral infarction, sICH – symptomatic intracerebral hemorrhage

There were no significant differences in the rates of favorable outcome at discharge as defined by an mRS≤ 2 or no change from baseline for those arriving with a mRS ≥3 (Table 2). However, mortality rates during the acute admission were doubled in patients with active cancer (28% vs. 14%, p=0.04).

**Table 2.** Outcomes

|  | All<br>(n=154) | Active<br>cancer<br>(n=84) | Cancer<br>history<br>(n=70) | p-value |
| --- | --- | --- | --- | --- |
| Favorable<br>recanalization (TICI<br>2b-3 %) | 132<br>(85%) | 72 (86%) | 60(86%) | 1 |
| Discharge Favorable<br>outcome (n=149, %) | 44 (29%) | 25 (32%) | 19 (27%) | 0.55 |
| Death during admission<br>(n=149, %) | 32 (21%) | 22 (28%) | 10 (14%) | 0.04 |
| 90days Favorable<br>outcome (n=139, %) | 55 (39%) | 24 (32%) | 30 (47%) | 0.08 |
| sICH (n=149, %) | 10 (7%) | 5 (6%) | 5 (8%) | 0.08 |
| Death at 90 days<br>(n=141, %) | 44 (34%) | 30 (38%) | 14 (21%) | 0.04 |

TICI – thrombolysis in cerebral infarction, sICH – symptomatic intracerebral hemorrhage

Similarly, favorable outcome at 90 days was seen in 32% of the patients with active cancer and in 47% of patients in the group of patients with a previous history of malignancy but no evidence of active disease (p=0.08). The 90 days mortality rates were significantly higher in the patients with active cancer (38% vs. 21%, p=0.04) and patients that survived were more often independent prior to stroke and more often reached target vessel recanalization (Supplementary Table 1).

We next compared between patients with favorable and unfavorable outcomes on day 90 after stroke. The results (Table 3) show that patients with favorable outcomes less often had diabetes, were more often independent at the time of stroke onset and more often reached favorable target vessel recanalization. On multivariable analysis (Table 4) that included variables that yielded a significance value of <0.1 on the univariate analyses, the presence of active cancer was not found as an independent predictor of favorable functional independence at 90days post stroke. In contrast being independent prior to stoke and achieving a favorable target vessel recanalization during EVT were positively associated with increased chances of achieving an independent functional state at 90 days post stroke.

**Table 3:** factors associated with favorable outcome

|  | Favorable<br>outcomes<br>N=55 | Unfavorable<br>outcomes<br>N=84 | p-value |
| --- | --- | --- | --- |
| Age, (±SD) | 72.5±12.2 | 73.4±12.1 | 0.68 |
| Sex, male (%) | 21 (38%) | 43 (51%) | 0.13 |
| Hypertension (%) | 30 (63%) | 58 (69%) | 0.08 |
| Diabetes (%) | 10 (18%) | 29 (34%) | 0.04 |
| Dyslipidemia (%) | 20 (36%) | 36 (43%) | 0.44 |
| Atrial fibrillation (%) | 27 (49%) | 30 (36%) | 0.12 |
| Smoking (%) | 11 (20%) | 26 (31%) | 0.15 |
| Ischemic heart disease (%) | 20 (36%) | 34 (41%) | 0.63 |
| Prior stroke (%) | 10 (18%) | 14 (17%) | 0.82 |
| Antiplatelets | 11 (20%) | 16 (19%) | 0.89 |
| Anticoagulants | 16 (29%) | 14 (17%) | 0.08 |
| Active cancer (%) | 24 (44%) | 50 (59%) | 0.08 |
| Vessel lesion (%) |  |  | 0.45 |
| Internal carotid | 6 (11%) | 16 (19%) |  |
| Middle cerebral | 36 (66%) | 49 (58%) |  |
| Tandem | 2 (4%) | 6 (7%) |  |
| Other | 11 (19%) | 13 (16%) |  |
| TOAST (%) |  |  | 0.10 |
| Cardioembolic | 31 (56%) | 33 (39%) |  |
| Large vessel atherosclerosis | 19 (35%) | 31 (37%) |  |
| Other known | 1 (2%) | 5 (6%) |  |
| Unknown | 4 (7%) | 15 (11%) |  |
| Baseline mRS (%) |  |  | 0.003 |
| 0-2 | 51 (93%) | 59 (71%) |  |
| 3 | 3 (5%) | 19 (23%) |  |
| 4 | 1 (2%) | 6 (6%) |  |
| Initial NIHSS score (±SD) | 14.6±5.7 | 15.6±5.9 | 0.34 |
| IV tPA (%) | 16 (29%) | 17 (20%) | 0.23 |
| TICI2b-3 (%) | 54 (98%) | 67 (80%) | 0.002 |
| Number of passes<br>(mean±SD) | 1.8±1.2 | 3.1±4.4 | 0.06 |
| sICH (%) | 1 (2%) | 9 (10%) | 0.05 |

mRS – modified Rankin Scale, NIHSS- National Institutes of Health Stroke Scale, TICI – thrombolysis in cerebral infarction, sICH – symptomatic intracerebral hemorrhage

**Table 4:** Multivariable regression for favorable outcomes at 90days post-stroke

| Variable |  | OR | 95% C.I. |  | p |
| --- | --- | --- | --- | --- | --- |
|  | Diabetes | 0.80 | 0.30 | 2.13 | 0.65 |
|  | Hypertension | 0.45 | 0.19 | 1.02 | 0.07 |
|  | On oral anticoagulant | 2.30 | 0.89 | 5.94 | 0.09 |
|  | Active cancer | 0.49 | 0.22 | 1.10 | 0.08 |
|  | Independent prior | 3.96 | 1.19 | 13.21 | 0.02 |
|  | TICI 2b-3 | 10.01 | 1.20 | 83.55 | 0.03 |
|  | sICH | 0.18 | 0.02 | 1.66 | 0.13 |

TICI – thrombolysis in cerebral infarction, sICH – symptomatic intracerebral hemorrhage

In contrast, on multivariable analysis the presence of active cancer remained an independent negative predictor of survival at 90 days post stroke (OR 0.41 95% CI 0.17-0.97) while favorable recanalization to TICI2b-3 (OR 3.66 95% CI 1.14-2.26) and being independent prior to stroke (OR 6.24 95% CI 2.26-17.22) were positively associated with survival (Table 5)

**Table 5:** Multivariable regression for survival at 90 days

|  |  | OR | 95% CI |  | p |
| --- | --- | --- | --- | --- | --- |
|  | TICI 2b-3 | 3.66 | 1.15 | 2.26 | 0.03 |
|  | active cancer | 0.41 | 0.17 | 0.97 | 0.04 |
|  | Independent status prior to stroke | 6.24 | 2.26 | 17.22 | <0.001 |
|  | tPA | 0.46 | 0.67 | 1.23 | 0.462 |
|  | Internal carotid occlusion | 2.56 | 0.80 | 13.37 | 0.47 |

TICI – thrombolysis in cerebral infarction,

## Discussion

The main findings of the current multi-center analysis are that outcomes tend to be less often favorable and mortality is significantly higher in patients with active cancer when compared with patients with a remote history of cancer. Importantly, stroke severity, occluded vessel location, favorable target vessel recanalization and sICH rates were similar among the groups and could not account for these findings. Patients with active caner were younger and also showed trends towards being more likely to be treated with bridging tPA as well as trends towards lower rates of having atrial fibrillation and large vessel atherothrombosis which were all expected to tilt mortality and favorable outcome rates towards better outcomes. However, mortality rates were significantly higher in active cancer patients and an active cancer status was associated with numerically lower rates of favorable outcome although this difference did not reach statistical significance.

Cancer patients are at increased risk for stroke, which may be related to a cancer-associated hypercoagulable state as well as to vessel wall damage secondary to radiation, biological and chemical treatments.^1-8^ Although the stroke risk decreases over time, it remains elevated even a decade following the cancer diagnosis. In addition, the prognosis is poorer in stroke patients with systemic malignancy than in those without.^21,28^

Patients with acute ischemic stroke and cancer can be treated with intravenous thrombolysis with rt-PA.^24^ Certain limitations such as bleeding tendencies, time from symptom onset and presence of large vessel occlusion may limit the use and effectiveness of intravenous thrombolysis. EVT is the treatment of choice for LVO stroke.^10^ Several studies have explored the safety and effectiveness of EVT in patients with malignancy.^12,14-20,22-24^ These studies were based on retrospective single center data with a low number of included patients^11,14-20^, on nationwide registry data that did not offer specifics about cancer type^22,24^ and only one prospective randomized trial.^23^ Data from the nationwide registries and the randomized controlled study showed that cancer patients comprise between 4.5% and 6.7% of all LVO patients treated with EVT which is slightly lower than the 12% seen in the current study. These differences may stem from different definitions of cancer and cancer in remission, which were used in different studies. Thus, we included patients with a history of remote cancer even years ago that were probably not included in cancer patients cohorts in some of the previous studies. Taken together, these studies recorded similar rates of favorable target vessel recanalization and symptomatic ICH^11,12,14,16-18,20,22-24^ but lower rates of favorable outcome and increased mortality rates in cancer patients when compared to patients without cancer.^11,12,16-18,20,22-24^ Only one prior study compared patients with active cancer to those with non-active cancer in remission.^24^ This is surprising in light of data suggesting that these populations may differ in stroke cause and distribution.^29^ The study^24^, which was based on national registry data in Korea showed that the outcomes of patients with active cancer have lower rates of favorable compared to those with cancer in remission that in turn resemble those of patients without cancer. The current data confirms these results and also shows higher mortality rates in patients with active cancer compared to those with remote cancer. Of note, mortality rates were higher in patients with active cancer both during the admission for the acute stroke and at 90 days after stroke. Most of the deaths that occurred between discharge and 90 days occurred in the active cancer group suggesting that at least some of the deaths observed in cancer patients at 90 days were related to the cancer itself and not to the stroke. However, since we did not have complete access to the causes of death we could not ascertain this hypothesis.

In these regards the current study presents novel data suggesting that patients with active cancer represent a group of high risk patients with especially poor outcome despite of achieving similarly high recanalization rates and low sICH rates compared with patients with a remote history of cancer. These data further suggest that at least some cancer patients including one third of those with active cancer and nearly half of the patients with remote cancer can benefit from EVT and have favorable outcomes.

The current study has several strengths. First, we are reporting on multi-center prospectively accrued data that reflects current everyday practice in centers that are EVT capable. As such, it provides much needed information for clinicians dealing with stroke in cancer patients at the emergency department level. Second, the included number of patients is relatively large and the frequency with which cancer associated LVO was observed is larger than that seen in larger nationwide registry data in the US or Korea.

Our study has limitations. First, although data collection was prospective, the data analysis was retrospective and therefore prone to bias. Second, we do not have information concerning cancer patients that were excluded from EVT because of reduced life expectancy or other family or physician based decisions. Third, we did not have access to causes of mortality in the included patients limiting our efforts to determine if mortality was secondary to cancer or cancer therapy or stroke related causes.

## Conclusion

Our study suggests that EVT is safe and efficacious in patients with history of malignancy at the time of stroke as well as in those with active cancer. While outcomes appear to be better in patients with a history of cancer, these results may suggest that all cancer patients with LVO may benefit from EVT despite higher mortality rates and lower favorable rates seen in patients with active cancer. Future studies identifying subpopulations of active cancer patients that may benefit more from EVT in contrast to those in which futile recanalization occurs are needed.

## Data Availability

Anonymized data can be shared upon request and in compliance with government rules

## Non-standard abbreviations and acronyms

EVT: endovascular thrombectomy
LVO: large vessel occlusion
mRS: Modified Rankin Scale
NIHSS: National Institutes of Health Stroke Scale
sICH: symptomatic intracerebral hemorrhage
TICI: thrombolysis in cerebral infarction

## Sources of Funding

This study was supported in part by an unrestricted grant from the Peritz and Chantal Scheinberg Cerebrovascular Research Fund

## Disclosures

FS, TS, AF, JM, ZIH, HH, SA, JMG and JEC have nothing to disclose

RRL received speaker honoraria from IscemaView, Boehringer Ingelheim, Pfizer, Jansen, Biogen, Medtronic and Abott and advisory board honoraria from Jansen.

## Supplemental materials

Table S1: Factors associated with mortality

## Notes

### Competing Interest Statement

The authors have declared no competing interest.

### Author Declarations

The study was approved at each individual center by the local institutional review boards at Hadassah Medical Organization, Tel Aviv Medical Center and Chaim Sheeba Medical Center and permissions to combine anonymized individual patient data into a unified dataset was granted with a waiver to obtain informed consent, given the retrospective nature of the data analysis.

