## Supplementary material for "Does malignancy status effect outcomes in patients with large vessel occlusion stroke and cancer that underwent endovascular thrombectomy?": Table S1

Supplementary Table 1: Factors associated with mortality

|  | Survived (n=102) | Deceased (n=44) |  |
| --- | --- | --- | --- |
| Age, (±SD) | 73.2±11.9 | 73.4±12.5 | 0.952 |
| Sex, male (%) | 46 (45%) | 21 (47%) | 0.770 |
| Hypertension (%) | 66 (65%) | 29 (66%) | 0.889 |
| Diabetes (%) | 27 (30%) | 17 (39%) | 0.142 |
| Dyslipidemia (%) | 42 (41%) | 22 (50%) | 0.324 |
| Atrial fibrillation (%) | 46 (45%) | 15 (34%) | 0.216 |
| Smoking (%) | 21 (21%) | 14 (32%) | 0.145 |
| Ischemic heart disease (%) | 37 (36%) | 17 (43%) | 0.431 |
| Prior stroke (%) | 19 (19%) | 7 (16%) | 0.694 |
| Antiplatelets | 23 (23%) | 7 (16%) | 0.362 |
| Anticoagulants | 23 (23%) | 7 (16%) | 0.362 |
| Active cancer (%) | 50 (49%) | 30 (68%) | 0.038 |
| Vessel lesion (%)  Internal carotid  Middle cerebral  Tandem  Other | 11 (11%)  61 (60%)  6 (6%)  21 (21%) | 11 (25%)  25 (57%)  2 (5%)  6 (13%) | 0.096 |
| Baseline mRS (%)  0-2 | 90 (91%) | 25 (58%) | 0.001 |
| Initial NIHSS score (±SD) | 14.9±5.8 | 15.5±6.4 | 0.578 |
| IV tPA (%) | 28 (28%) | 6 (14%) | 0.070 |
| TICI2b-3 (%) | 91 (92%) | 33 (75%) | 0.006 |
| Number of passes (mean±SD) | 2.6±3.9 | 2.4±1.9 | 0.836 |
| sICH (%) | 6 (4%) | 4 (9%) | 0.488 |

mRS – modified Rankin Scale, NIHSS – National Institutes of Health Stroke Scale, tPA – tissue plasminogen activator, TICI – Thrombolysis in Cerebral Infarction, sICH – symptomatic intracranial hemorrhage
